# A simple ecological model captures the transmission pattern of the coronavirus COVID-19 outbreak in China

**DOI:** 10.1101/2020.02.27.20028928

**Authors:** Feng Zhang, Jinmei Zhang, Menglan Cao, Cang Hui

## Abstract

The rapid spread of the 2019 novel coronavirus (COVID-19), initially reported in the city of Wuhan in China, and quickly transmitted to the entire nation and beyond, has become an international public health emergency. Estimating the final number of infection cases and the turning point (time with the fastest spreading rate) is crucial to assessing and improving the national and international control measures currently being applied. In this paper we develop a simple model based on infectious growth with a time-varying infection rate, and estimate the final number of infections and the turning point using data updated daily from 3 February 2020, when China escalated its initial public health measures, to 10 February. Our model provides an extremely good fit to the existing data and therefore a reasonable estimate of the time-varying infection rate that has largely captured the transmission pattern of this epidemic outbreak. Our estimation suggests that (i) the final number of infections in China could reach 78,000 with an upper 95% confidence limit of 88,880; (ii) the turning point of the fastest spread was on the 4^th^ or the 5^th^ of February; and (iii) the projected period for the end of the outbreak (i.e., when 95% of the final predicted number of infection is reached) will be the 24^th^ of February, with an upper 95% confidence limit on the 19^th^ of March. This suggests that the current control measures in China are excellent, and more than sufficient to contain the spread of this highly infectious novel coronavirus, and that the application of such measures could be considered internationally for the global control of this outbreak.

## Introduction

The Wuhan coronavirus (COVID-19) possibly originated from bats^1^ and caused the current pneumonia outbreak, raging since December 2019. As the COVID-19 has an excessively high human-to-human transmission capability, and the early outbreak stage coincided with the travel rush of the Chinese Spring Festival, the pandemic has been spreading rapidly^2^. According to reports by the National Health Commission of China (NHCC), the first confirmed case was reported in Wuhan (Hubei Province) on 8 December 2019; the following day there were also confirmed cases in the major Chinese cities of Beijing and Shanghai, as well as in the neighbouring countries of Japan, Thailand and South Korea; as of 25 January, a total of all 31 Chinese provinces, as well as Hong Kong, Macau, Taiwan, the United States, France and Australia reported pandemic cases of COVID-19. It is apparent that the pandemic is still ongoing both in China and internationally.

In the meantime, Chinese authorities have taken prompt public health measures to control this outbreak. Medical teams and supplies from all over the country flooded into Wuhan. All major cities in Hubei Province were sealed off to contain the spread of the virus, with quarantine imposed on all people exposed to the virus. China has striven to limit the spread of the virus through prohibiting human traffic and movement at an unprecedented level. The World Health Organization (WHO) has also taken a series of measures to consolidate global scientific research, quarantine, and surveillance efforts to help fight the pandemic. At present, WHO is working with partner agencies to enhance the global diagnostic capabilities of COVID-19, improve surveillance and the tracking of disease transmission, along with mobilizing global powers to coordinate research on vaccines, therapies and diagnostic technologies for the virus.

For effective monitoring and containment of the pandemic, it is therefore crucial to predict and estimate the spreading dynamics of this novel infectious disease from a modelling point of view. Much has been done using the traditional SIR epidemiological models^3-5^. However, such models often suffer from unreliable model structure and parameterization; these problems are especially pronounced in this case when the pathology and the transmission pathways of the virus remain unclear, making rapid, accurate forecasting impossible. Here we propose a population ecology model with a time-varying infection rate that does not rely on the detailed pathology of the disease but could accurately capture the spreading dynamics of the virus. This model allows us to estimate and predict the spreading dynamics of the COVID-19 with reliable accuracy, given the current healthcare measures in place.

## Materials and Methods

We first develop a simple population dynamic model with a time-varying parameter. Let *N*(*t*) be the number of infected cases, with its temporal dynamics following an ordinary differential equation, *dN*(*t*)/*dt* = *r*(*t*)*N*(*t*), where *r*(*t*) is the time-dependent infection rate. This model applies when the susceptible population is large but the outbreak is limited and temporary so that the demographic dynamics of the entire population remain unchanged (i.e., the additional death and recovery rates caused by the infection among all residents of the Hubei Province are negligible). With this simple model, we can estimate the turning point as the moment when the number of infected cases increases at the fastest speed, happening when *d*^2^ *N*(*t*)/*dt*^2^ = 0. That is, the turning point is the solution to the equation *dr*(*t*)/*dt* + *r*(*t*)^2^ = 0. Notably, *dr*(*t*)/*dt* < 0 is necessary condition for existing turning point. In practice, the infection rate can be estimated as *r*(*t* + 1/2) = ln(*N*(*t* + 1)) - ln(*N*(*t*)), where *t* is measured in days.

We compiled the daily numbers of diagnosed COVID-19 infections from 10 January to 10 February 2020 in Wuhan city, Hubei Province, and the whole of China from the website of the NHCC (www.nhc.gov.cn) and the Health Commission of Hubei Province (wjw.hubei.gov.cn). This allowed us to analyse the data at four levels: Wuhan city, Hubei Province, the rest of China, and the whole of China. As there were two large-scale control measures implemented on 26 January and 2 February, we fitted the time-dependent infection rate *r*(*t*) with exponential functions for two periods: from 27 January to 2 February, and from 3 February to 10 February. We estimated the trajectories of the cumulative number of infected cases using the fitted infection rate according to the difference equation, *N*(*t* + *τ*) = exp(*r*(*t* + *τ*/2)*τ*)*N*(*t*), with *τ* = 0.05 (day). The daily increase of infected cases can be calculated as (*N*(*t* + *τ*) - *N*(*t*))/*τ*, and the turning point can be identified according to the condition *dr*(*t*)/*dt* + *r*(*t*)^2^ = 0.

## Results and Discussions

Since 26 January when China implemented strict nationwide control measures, a clear pattern of transmission has emerged. Albeit minor declines can be observed, the time-varying rate of infection has dropped exponentially for the whole of China and Hubei Province, although the rate still increases in Wuhan city (red lines in Fig.1). This means that the first nationwide control measure (implemented on 26 January) failed to contain the outbreak and the spread remained out of control.

**Fig. 1:**
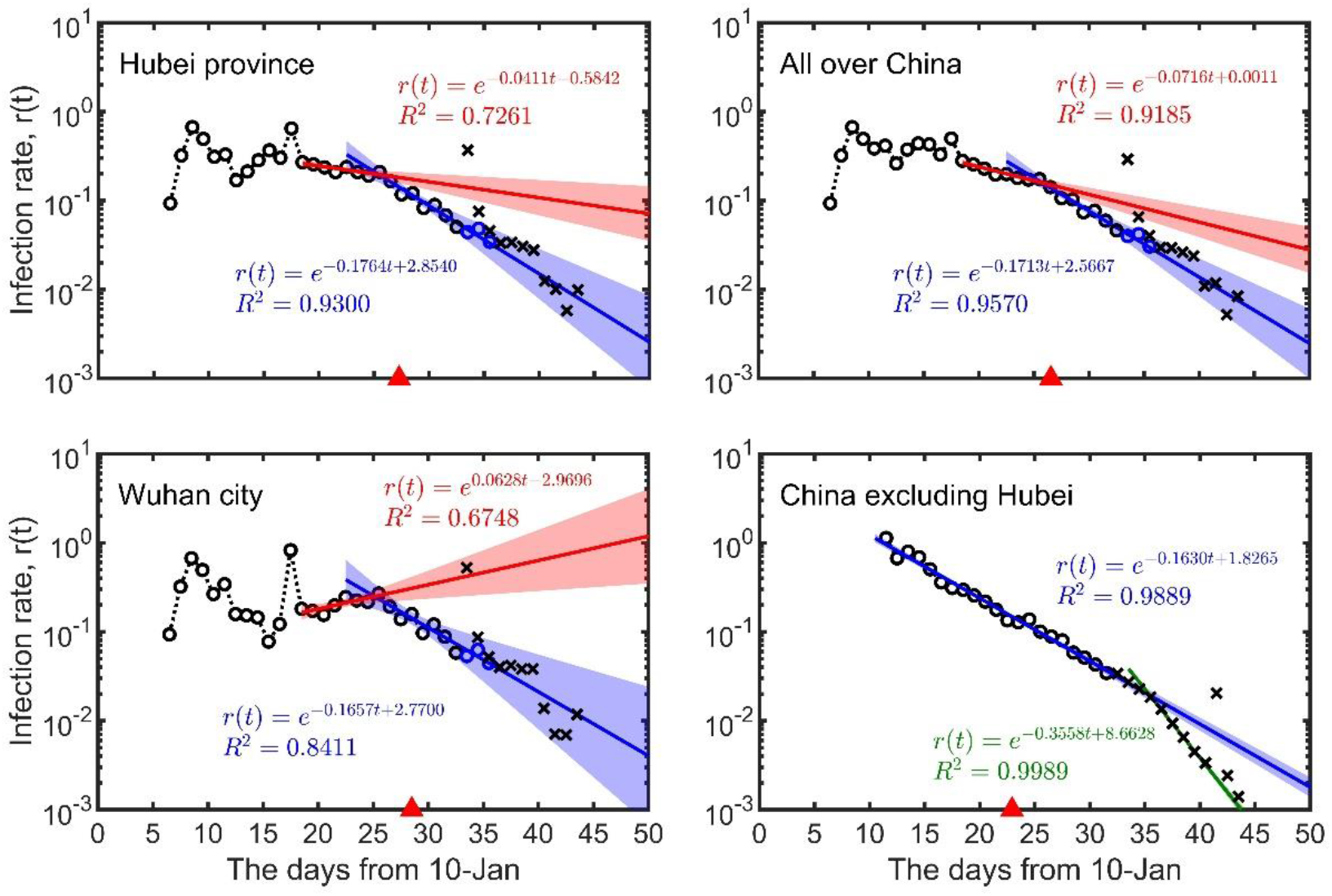
The time-dependent infection rate. Points show data reported by the National Health Commission of China and the Health Commission of Hubei Province. Red lines and blue lines are from the regression of the data from 27-Jan to 2-Feb and from 3-Feb to 10-Feb, respectively. Green line represents the regression of 13-Feb to 18-Feb. Shadow belts indicate 95% confidence intervals. Open circles are the data before 10-Feb. Black crosses represent after predictions (the data from 12-Feb to 14-Feb include clinical diagnoses, instead of nucleic acid detection alone, where blue circles represent data excluding clinical diagnoses). Red triangles indicate predicted turning points.

According to the data from 3 February to 10 February, the infection rates of Wuhan city, Hubei Province and the whole of China were all declining at largely the same rate (a decrement of 0.16-0.18 per day; the slope of blue lines in Fig.1), similar to the rate of decline in the rest of China when the Wuhan and Hubei cases were excluded (the slope of the blue curve in Fig.1). It is worth pointing out that the infection rate has declined at a steady pace for areas outside Hubei Province. This suggests that the second nationwide control measure (implemented on 2 February) was effective. The fitted model from 3 February to 10 February can accurately predict the daily and accumulated infection numbers on 11 February. However, on 12 February, Hubei Province changed its clinical diagnostic approach, leading to a sharp rise of diagnosed cases and a reported daily increase of 15,151 infections; this unforeseeable change made the number of reported cases much greater than model predictions. Nevertheless, the reports after 12 February suggest that the change of clinical diagnostic approach did not affect the transmission pattern, and the daily infection rate reverted to our model prediction (black crosses in Fig.1). As such, we have estimated the spreading dynamics after 13 February according to our fitted model (from 3 February to 10 February). Our results show that the total numbers of infections in Wuhan city, Hubei province, and the whole of China, are 48,530, 64,640 and 77,990 respectively (Table 1). By subtraction, this means a total number of 13,350 for the rest of China excluding Hubei Province, which is highly consistent with the model prediction of 13,660 (see Table 1).

**Table 1:** Predictions for the coronavirus COVID-19 outbreak in China (time refers to the number of days from 10-Jan; numbers in parentheses represent 95% confidence intervals).

| Region | Turning point (day) | Final infections (Thousand) | Day of 95% prediction |
| --- | --- | --- | --- |
| Wuhan city | 28.51 | 48,530 (41,360 – 95,450) | 45.47 (38.14, 69.79) |
| Hubei Province | 27.27 | 64,640 (59,240 – 79,880) | 42.85 (38.19, 52.60) |
| Entire China | 26.54 | 77,990 (72,980 – 88,880) | 42.62 (38.80, 49.34) |
| China excl. Hubei | 22.93 | 13,660 (11,810 – 16,070) | 37.07 (35.72, 38.83) |

Our model also suggests that the spreading dynamics follow a typical S-shaped logistic curve. This means that there is a turning point when the rate of infection has reached its peak. Transmission accelerates when approaching the turning point, then decelerates after the turning point. Our model indicates a turning point around the 4^th^ to the 5^th^ of February (Fig.1 & 2). The duration of the outbreak, for reaching 95% of the final predicted number of infection from 10 January, was estimated to last 46 days in Wuhan city, 43 days in Hubei and the whole of China, and 37 days when Hubei was excluded (Table 1); it was estimated the outbreak would subside at the end of February in China.

**Fig. 2:**
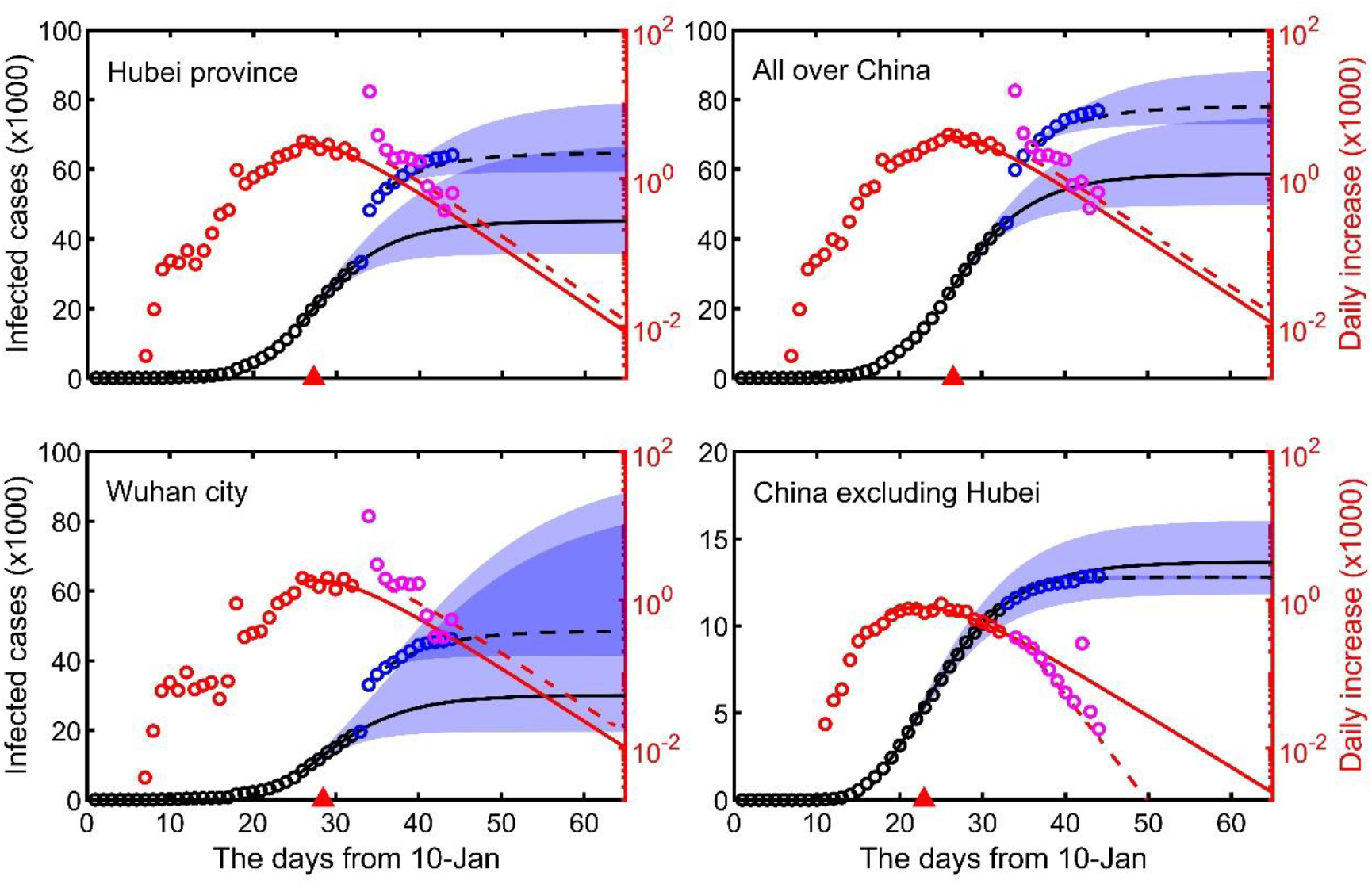
The predicted dynamics of infection over time. Solid lines represent the predictions based on the models fitted to the data from 3-Jan to 10-Jan, while dashed lines represent revised predictions due to the change of diagnostic method from 12-Feb. Shadow belts represent a 95% confidence interval. Black and red points represent the data before 10 February (for model fitting), while blue and pink points represent the data afterwards. Red triangles indicate predicted turning points.

Our model assumed an infinite size of a susceptible population trivially affected by infection, recovery and control. This makes our model different from the traditional SIR or SEIR epidemiological models, where the rise of infection cases is due to the large susceptible population, while the decline of infection cases is due to the depletion of the susceptible population. Such typical pattern of epidemiological models is obviously unsuitable for the spread of COVID-19 in China with susceptible populations of such massive size (e.g., >11 million residents in Wuhan alone and ∼60 million residents in Hubei). Epidemiological models could be more suitable for modelling the spread of COVID-19 among the 3,400 passengers and crew members on the *Diamond Princess* cruise ship currently docked in Japan’s Yokohama harbour for quarantine.

Our simple model captured the dynamic nature of the infection rate, which can be adjusted flexibly according to updated data. This allows us to predict short-term spreading dynamics with accuracy, which is essential for the live monitoring of the epidemic. According to the time dependency of the infection rate, we could also assess the effectiveness of implemented control measures and provide modelling support to adjust any public health measures. Our results showed three stages of the COVID-19 spreading: (1) the early stage with no clear dynamic patterns, due largely to lack of awareness and diagnostic capability; (2) from 27 January to 2 February there were nationwide control measures but with no clear effect on the control of the disease, especially in Wuhan city; (3) since 2 February another coordinated nationwide control measure was implemented, with the clear effects of containing the spread of the virus, decelerating and eventually reducing the epidemic. Lessons learnt from these nationwide control measures applied after 2 February should be communicated to the WHO, to help contain the spread of COVID-19 worldwide.

## Data Availability

The data can be obtained directly from the CDCC reports.

## Acknowledgement

FZ is supported by National Natural Science Foundation of China (No.31360104) and Anhui University (S020118002/101); CH is supported by the National Research Foundation of South Africa (grant 89967).

## References

1. Zhou, P. et al. A pneumonia outbreak associated with a new coronavirus of probable bat origin. Nature, doi:10.1038/s41586-020-2012-7 (2020).

2. Li, Q. et al. Early Transmission Dynamics in Wuhan, China, of Novel Coronavirus-Infected Pneumonia. N Engl J Med, doi:10.1056/NEJMoa2001316 (2020).

3. Wu, J. T., Leung, K. & Leung, G. M. Nowcasting and forecasting the potential domestic and international spread of the 2019-nCoV outbreak originating in Wuhan, China: a modelling study. Lancet, doi:10.1016/S0140-6736(20)30260-9 (2020).

4. Tang, B. et al. Estimation of the Transmission Risk of the 2019-nCoV and Its Implication for Public Health Interventions. J Clin Med 9, doi:10.3390/jcm9020462 (2020).

5. Keeling, M. J. & Rohani, P. Modeling Infectious Diseases. (Princeton University Press, 2008).

